# Factors associated with osteoarthritis in elderly people in Shanghai: A cross-sectional study

**DOI:** 10.1101/2023.09.28.23296309

**Authors:** Xueyao Liang, Bo Zheng, Xiyou Cao, Shuangfei Xu, Qiongmei Zhang, Qing Fei, Nanqian Lu, Kai Zhang, Qingchun Liu, Ying Han, Xiaoyin Hu, Weibing Wang, Bing Liu, Mengfan Li, Ying Wu, Yefeng Zhao

## Abstract

**Background:** Osteoarthritis (OA) is a degenerative disease of mostly’ middle-aged and elderly people that contributes a high burden of disease in China and worldwide. However, many putative risk factors associated with OA in elderly populations remain substantially unclear.

**Methods:** In our cross-sectional study, 2177 residents of the Putuo District of Shanghai aged 40 years and above were recruited to complete a questionnaire that queried basic health information and assessed the prevalence of OA. A non-conditional and an ordinal logistic regression model was used to analyze the associations between putative risk factors and the severity of confirmed OA.

**Results:** Among 2099 eligible participants, the average age was 65.21 years (41.4% were male), and 551 self-reported OA [prevalence = 26.25% (24.13, 28.51)]. The prevalence of knee OA were the most common (23.11%, 21.12 to 25.23). OA was associated with age ≥ 60 years (aOR = 1.56; 1.18, 2.07), overweight BMI (24∼27.9 kg/m2) (1.30; 1.02 to 1.65), female sex (1.66; 1.30 to 2.12), unmarried status (1.41; 1.03 to 1.90), a family history of OA (6.23; 4.17 to 9.38), two or more comorbidities (2.18; 1.52 to 3.11), and prior injury to any joint (12.02; 4.86 to 31.22).

**Conclusions:** Old age, Female sex,family history and multiple comorbidities were significant contributors to promote the severity of OA. Appropriate prevention and treatment strategies must be developed to reduce the burden of the disease.

## 1 INTRODUCTION

OA is the most common degenerative disease of joints [1] caused by fibrosis, chapping, ulceration, and attrition of articular cartilage, with clinical manifestations of joint pain, tenderness, limitation of movement and articulation, occasional effusion, and local inflammation of varying degrees[2]. As a commonly irreversible disease of middle-aged and elderly (aged 40 years old and above)[3], OA is part of the aging process, which will seriously affect the patient’s quality of life. The final disability rate of this disease is 53%, and advanced knee OA can lead to the need for joint-replacement surgery[1, 4, 5]. OA is recognized as a major public health challenge; it affects an estimated 32 million people in the US and more than 240 million people worldwide[2]. Globally, the age-standardized point prevalence, annual incidence rate, and age-standard YLD rate of OA were increasing from 1990 to 2017[6].

OA similarly contributes to a high burden of disease in China[7]; it is estimated that around 26.1 million individuals in China had OA in 1990, and this number rose to 61.2 million in 2017[8]. The prevalence of OA in China varies according to sociodemographic, economic, and geographic factors. Xu Tang’s research showed that the North and East regions of China had the lowest prevalence of symptomatic knee OA (5.4% and 5.5%, respectively), followed by the Northeast (7.0%), South-Central (7.8%), and Northwest (10.8%) regions, with the highest prevalence seen in the Southwest region (13.7%)[9]. According to the Shanghai Bureau of Statistics[10], in 2021, the city’s registered population aged 60 years and over was about 5,324,100; approximately 366,000 of these individuals were in Putuo District, accounting for 40.99% of the total population in the district. OA is a major public health concern that results in decreased quality of life among middle-aged and older adults[11]. Therefore, epidemiologic studies aimed at identifying risk factors that contribute to OA in China are needed, as are explorations of prevention and treatment strategies appropriate for older individuals.

Previous studies found that OA is related to many factors, such as age, gender, and region[9, 12, 13]. OA prevalence and YLDs are higher in females than in males, and the YLD rate increases with age[8]. While a multitude of factors contribute to the development of OA, many putative risk factors associated with OA in elderly populations remain substantially unclear, except for age, sex, and BMI. Additionally, the associations between putative risk factors and OA severity are uncertain. Given society’s responsibility to protect the elderly population, it is essential that researchers identify the risk factors associated with musculoskeletal disease in later life. This study aimed to investigate the prevalence of OA in a community of elderly people in the Putuo District of Shanghai, and to determine putative risk factors associated with confirmed OA and the severity thereof.

## 2 Methods

### Study design and setting

This study was cross-sectional and involved distributing a survey to collect information on factors potentially associated with OA, as well as demographics, lifestyle, past medical history, general health, occupational history, and participation in sports and physical activity. Our trained investigator introduced the purpose of the study and provided information to participants on filling out the questionnaire. Our survey was carried out between Oct 2020 and Mar 2021 in the Putuo District of Shanghai. Implied consent to participate was obtained from all participants completing the study questionnaire. The study was approved by the Ethics Committee of the Shanghai Putuo Liqun Hospital (No. RT-201911).

### Sampling and eligibility criteria

The recruitment of eligible individuals was performed using multi-stage stratified sampling. Lists of hospitals were obtained with the help of workers from Putuo District, and the Liqun Hospital was chosen. Sufficient clinical departments were sampled from the list of all clinical departments in the hospital, and eligible patients of the chosen departments were contacted in sequence. A trained investigator familiar with the purpose of the study provided information on filling out the questionnaire.

A signed informed consent form was obtained from all participants included in the study. All included participants had lived in the Putuo District for at least 6 months prior to being enrolled in the study. Inclusion criteria for participants were aged 40 years and older. The exclusion criteria for this study were: 1) the presence of a severe mental or neurological disorder; 2) the presence of rheumatoid arthritis, reactive arthritis, ankylosing spondylitis, or another secondary joint disease; and 3) data missing on the a priori confounders of age, sex, and BMI.

### Survey

The questionnaire consisted of four sections: demographic information, health status, living/working environment and dietary conditions and physical examination. The Western Ontario and McMaster Universities OA Index (WOMAC index, Chinese version)[14] was additional part that was given only to those participants with confirmed OA (self-reported had been diagnosed OA by X-rays or labs tests). Demographic information, living/working environment and dietary condition referred to (as examples) the respondent’s age, sex, height, weight, residence, educational level, working posture, and per capita annual household income information. The obtained health status information comprised information on medication history, physical exercise, comorbidities, joint pain, trauma history, surgery history, OA history of family members, etc. The WOMAC is a self-assessment questionnaire comprising 24 items on three subscales: pain (5 items), stiffness (2 items), and physical function (17 items)[15]. The overall WOMAC score (index) is determined by summing the scores across the three dimensions; the score can range from 0 to 240[15]: <80 indicates mild OA, 80-120 is moderate OA, and >120 represents severe OA. The Chinese version of WOMAC has strong internal consistency (Cronbach’s alpha = 0.840.96) and acceptable test-retest reliability[16]. For the WOMAC index, our study achieved a Cronbach’s alpha score of 0.986. In this study, symptomatic OA was adopted as the diagnostic criteria, which refers to the presence of recurring joint pain and other symptoms, including joint pain, stiffness and limited mobility, in the past month on the basis of imaging OA

### Statistical analysis

The collected data from questionnaires were entered with Epidata version 3.4 for data management and to be proofread. Mean (95% CI) or median/interquartile ranges were used to describe the distribution of continuous variables, and the frequency and component ratio were used to describe the characteristics of categorical variables. The Mann-Whitney U test was used to compare means and the Chi-square test was used to compare rates.

Univariate logistic regression was used to analyze crude associations between each independent variable and confirmed OA, which was reported as a crude odds ratio (cOR) with 95% confidence. All factors were entered separately into Adjusted model 1 and adjusted for the a priori confounders of age, sex, and BMI. The obtained Adjusted model 2 was then fitted to the a priori confounders plus any significant factors/variables identified in Adjusted model 1. Adjusted model 2 involved the stepwise multivariate analysis of any significant factors/variables identified in Adjusted 1 model plus the a priori confounders. Meanwhile, we used univariate ordinal logistic regression to determine the univariate associations between significant factors identified for confirmed OA and the severity of OA. All significant factors (P < 0.05) were included in our multivariate analysis. P values and 95% CIs were used to assess statistical significance in all models. We conducted all analyses in R.4.1.1 software (Foundation for Statistical Computing, Vienna, Austria. ISBN 3-900051-07-0, URL https://www.r-project.org/). Statistical significance was set at a two-side p value < 0.05.

## 3 Results

### Demographic, lifestyle, and health characteristics of participants

A total of 2177 questionnaires were returned between Oct 2020 and Mar 2021. (details of excluded participants are available in online supplemental Figure 1). Of those included in the analysis(n = 2099), the mean age was 65.21 years (95% CI: 64.79, 65.63), 58.6% were female (1230/2099), and the average BMI were 24.85(24.42-25.27). The average cumulative working years was 29.03 years (28.65, 29.41), and 33.4% (702/2099) of respondents walked for their daily travel. The percentage reporting married status was significantly higher among male participants (88.8%) than female participants (85.4%) (P < 0.001). Of the participants, 82.9% (1020/1230) of females took stairs during their daily or work life, while only 77.6% (674/869) of males reported habitual stair use. A family history of OA was reported by 4.8% (42/869) of male participants and 8.5% (105/1230) of female participants; thus, significantly more female participants had a family history of OA compared to male participants (P < 0.001). Among all participants, 971 (46.3%) had self-reported comorbidities and 655 (31.2%) were receiving long-term any medical treatment (see Table 1).

**Table 1:** Anthropometry, Lifestyle, and Health Factors.

| Characteristics | All<br>(n = 2099) | Male<br>(n = 869) | Female<br>(n = 1230) | *P*<br>Value |
| --- | --- | --- | --- | --- |
|  | Mean (95% CI) | Mean (95% CI) | Mean (95% CI) |  |
| Age, years | 65.21 (64.79 ,65.63) | 65.69 (65.05, 66.34) | 64.87 (64.32, 65.42) | 0.057 |
| Height, cm | 163.98 (163.67 ,164.29) | 169.92 (169.52, 170.32) | 159.78 (159.51, 160.05) | **< 0.001** |
| Weight, kg | 71.23 (69.59, 72.87) | 63.75 (62.25, 65.24) | 66.85 (65.73 ,67.97) | **< 0.001** |
| Body mass index, kg/m<sup>2</sup> | 24.85 (24.42 ,25.27) | 24.66 (24.09, 25.23) | 24.98 (24.38, 25.58) | 0.453 |
| Duration of working career, years | 29.03 (28.65 ,29.41) | 32.02 (31.36 ,32.67) | 26.9 (26.49 ,27.31) | **< 0.001** |
| Ethnic Han, % | 1879 (89.5) | 796 (91.6) | 1083 (88.0) | **0.011** |
| Married, % | 1823 (86.9) | 772 (88.8) | 1051 (85.4) | **0.028** |
| Junior high school and below, % | 889 (42.4) | 354 (40.7) | 535 (43.5) | 0.224 |
| Household income per capita <5000 CNY, % | 1314 (62.6) | 534 (61.4) | 780 (63.4) | 0.088 |
| Medical insurance for urban workers, % | 1541 (73.4) | 606 (69.7) | 935 (76.0) | **0.002** |
| 0-3 hours/per week for physical exercise, % | 1108 (52.8) | 468 (53.9) | 640 (52.0) | 0.626 |
| Both sitting or standing in work, % | 759 (36.2) | 297 (34.2) | 462 (37.6) | 0.186 |
| Ambidextrous, % | 207 (9.9) | 74 (8.5) | 133 (10.8) | 0.119 |
| Walk for daily travel, % | 702 (33.4) | 193 (22.2) | 509 (41.4) | **<0.001** |
| Taking stairs for daily life or work, % | 1694 (80.7) | 674 (77.6) | 1020 (82.9) | **0.003** |
| Balanced diet, % | 1477 (70.4) | 590 (67.9) | 887 (72.1) | 0.082 |
| Family history of OA, % | 147 (7.0) | 42 (4.8) | 105 (8.5) | **0.004** |
| Any history of surgery, % | 98 (4.7) | 32 (3.7) | 66 (5.4) | 0.183 |
| Any long-term medical treatment*, % | 655 (31.2) | 250 (28.8) | 405 (32.9) | 0.128 |
| Any current disease, % | 971 (46.3) | 390 (44.9) | 581 (47.2) | 0.307 |

Data are presented as means with 95% confidence intervals (95% CIs) or as proportions (%). The P values represent comparison between male and female participants using unpaired t-test or Chi-square analysis. Statistically significant differences are bolded.
\*Chronic or intermittent use over 6 months.

### Prevalence of OA and OA-related pain

Of participants, 551 were identified as having confirmed OA [prevalence = 26.25% (95% CI: 24.13, 28.51)]; more women (70.4%) than men (29.6%). Knee OA had the highest prevalence (23.11%) followed by interphalangeal OA (2.38%), hip OA (1.95%), and other-region OA (1.00%). For the prevalence of recurrent joint pain in the prior month, 20.68% (18.80, 22.69), 7.05% (5.98, 8.26), 6.34% (5.33, 7.48), and 4.67% (3.81, 5.66) of participants reported such pain in the knee joint, interphalangeal joint, hip joint, and other joints, respectively. Among those reporting pain of any joint in the prior month, 73.2% (595/813) were female and 26.8% (218/813) were male (see Figure 2).

**Figure 1.**
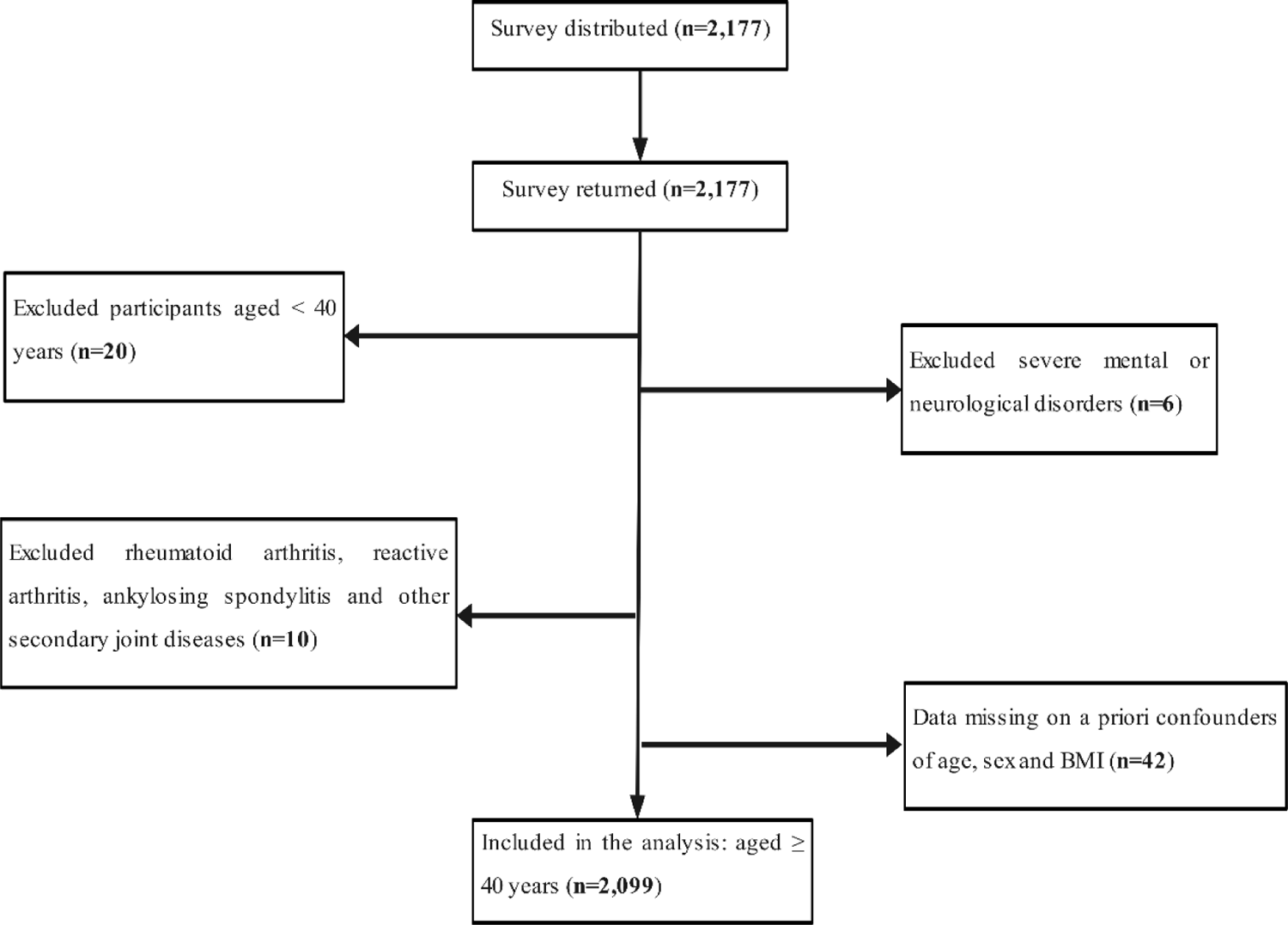
Flow chart depicting the number of participants included in this study.

**Figure 2.**
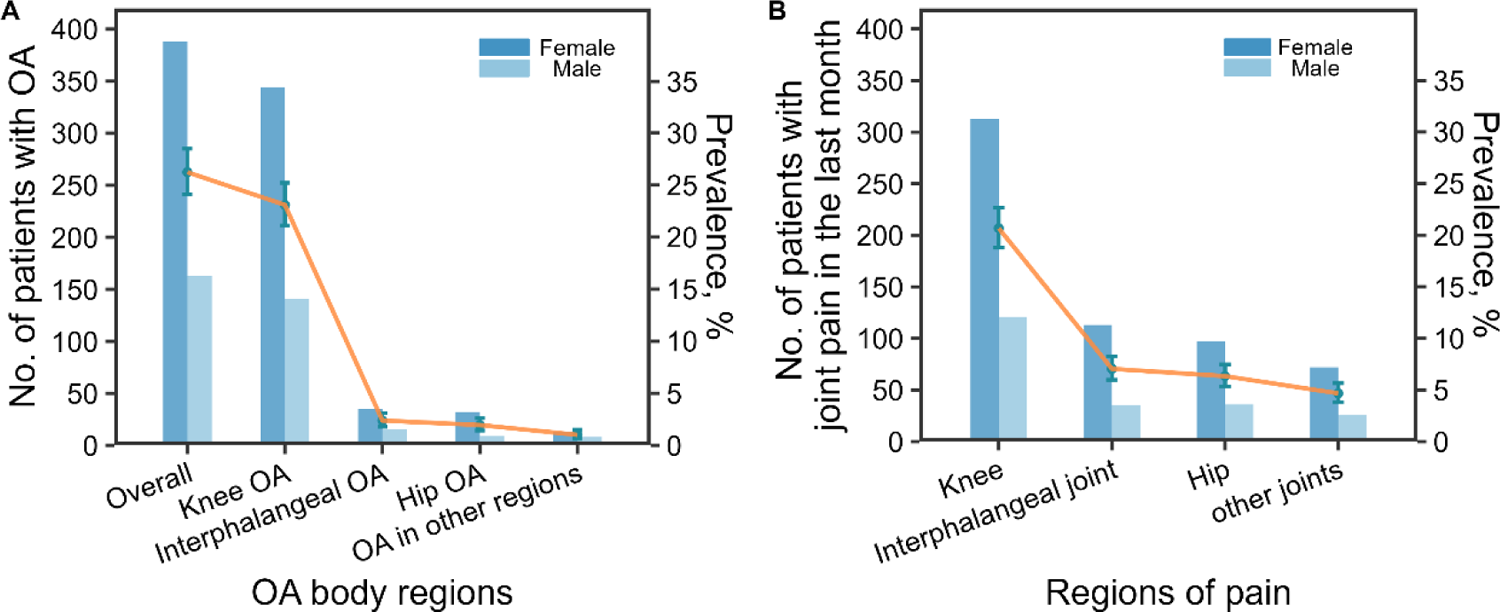
Prevalence of clinically confirmed OA (A) and recurrent joint pain in the last month (B) among different body regions.

### Factors associated with OA

In our ultimate stepwise multi-variable (Adjusted model 2) analysis of all significant factors/variables from Adjusted model 1 (Supplementary Table 2) plus the a priori confounders revealed that OA was associated with age ≥ 60 (aOR = 1.56; 1.18, 2.07, P < 0.001), overweight BMI (24∼27.9 kg/m^2^) (aOR = 1.30; 1.02, 1.65, P = 0.040), female sex (aOR = 1.66; 1.30, 2.12, P < 0.001), unmarried status, divorced or widowed (aOR = 1.41; 1.03, 1.90, P = 0.030), a family history of OA (aOR = 6.23; 4.17, 9.38, P < 0.001), two or more comorbidities (aOR = 2.18; 1.52, 3.11, P < 0.001), and prior injury of any joint (aOR = 12.02; 4.86, 31.22, P < 0.001). Similarly, we identified potential protective factors [e.g., professional and technical work (aOR = 0.51; 0.29, 0.87, P = 0.010), ambidextrous (aOR = 0.47; 0.23, 0.97, P = 0.040), use of bicycle for daily travel (aOR = 0.61; 0.44, 0.83, P < 0.001), use of public transportation or other mode for daily travel (aOR = 0.59; 0.45, 0.77, P < 0.001)] in our ultimate stepwise multi-variable analysis (see Figure 3).

**Figure 3.**
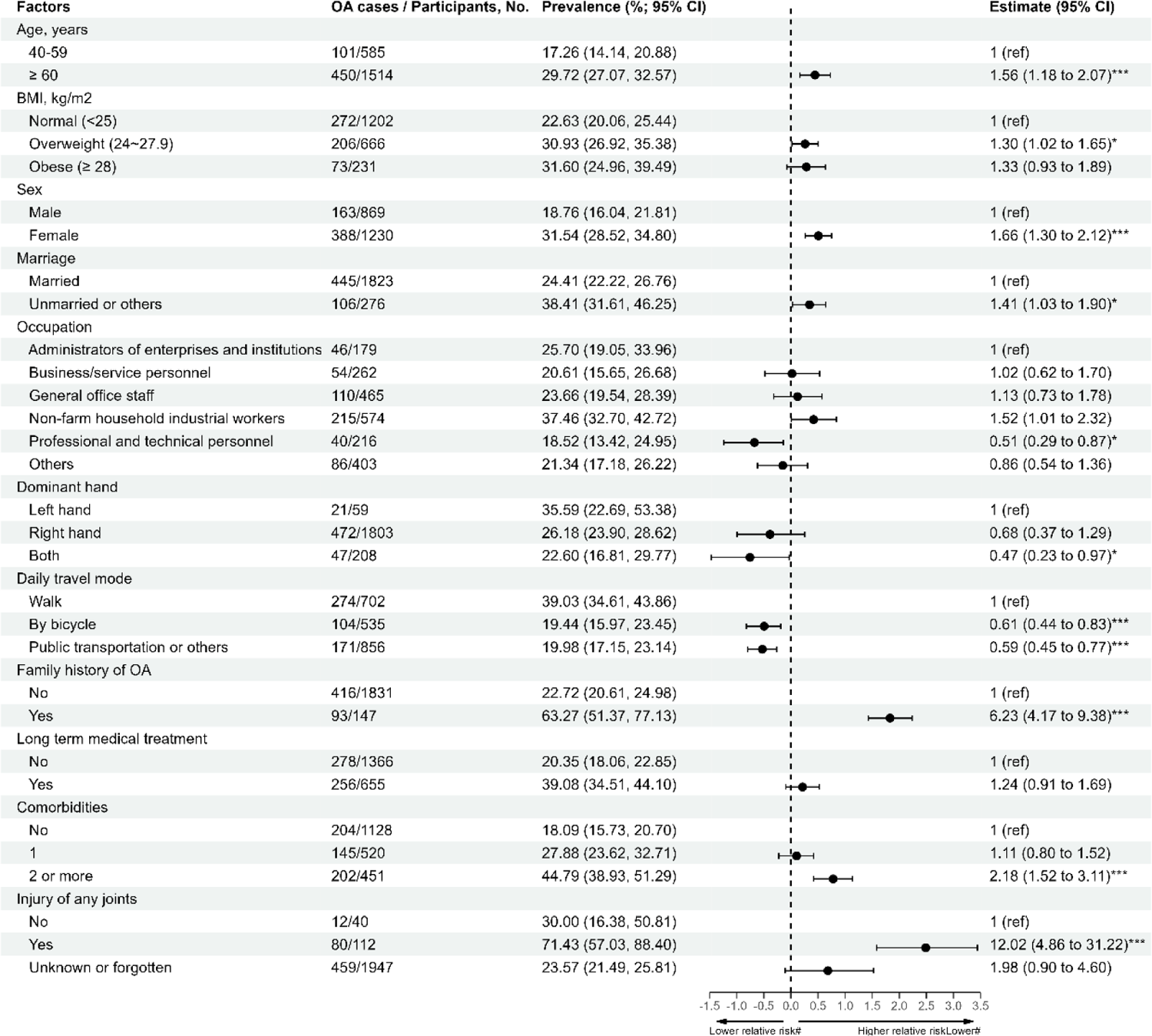
Demographic, lifestyle, and biomechanical factors of OA. Stepwise multivariable logistic regression analysis of significant factors/variables identified in Adjusted model 1 (Supplementary Table 2) plus a priori confounders of age, sex, and BMI. The error bar and dots represent the adjusted relative risk of OA with a logarithmic scale. The error bars indicate 95% CIs. The vertical dotted line at 0.0 is a reference for the x-axis. *P < 0.05, **P < 0.01, ***P < 0.001; ^#^ logarithmic scale.

### Factors associated with the severity of OA

Of participants with confirmed OA, the overall response rate to the WOMAC questionnaire was 71.9% (396/551); of these respondents, 367 scored as mild OA (92.7%), 11 as moderate OA (2.8%), and 18 as severe OA (4.5%). In our univariate ordinal logistic regression, the use of a bicycle for daily travel (cOR = 0.12; 95% CI: 0.02, 0.95) and non-long-term medical treatment tended to be associated with less severe OA compared to walking for daily travel and long-term medical treatment (cOR = 4.00; 1.57, 10.19), respectively; however, these differences did not reach the level of significance (P _by bicycle_ = 0.234; P _long-term medical treatment_ = 0.626) in multivariate analysis. Unmarried or other (divorced/widowed) status participants had significantly more severe OA than those of Married (cOR = 4.97, 2.28, 10.84; aOR = 3.75, 1.53, 9.19, P = 0.004). Participants reporting a family history of OA (cOR = 3.09, 1.30, 7.34; aOR = 4.73, 1.73, 12.91, P = 0.002) tended to have significantly more severe OA. Our results also showed that an increasing number of comorbidities contributed to increasing the severity of OA, with significant contributions observed for participants with two or more comorbidities (cOR = 7.91, 2.30, 27.16; aOR = 8.53, 1.75, 41.54, P = 0.008) (see Table 2).

**Table 2.**
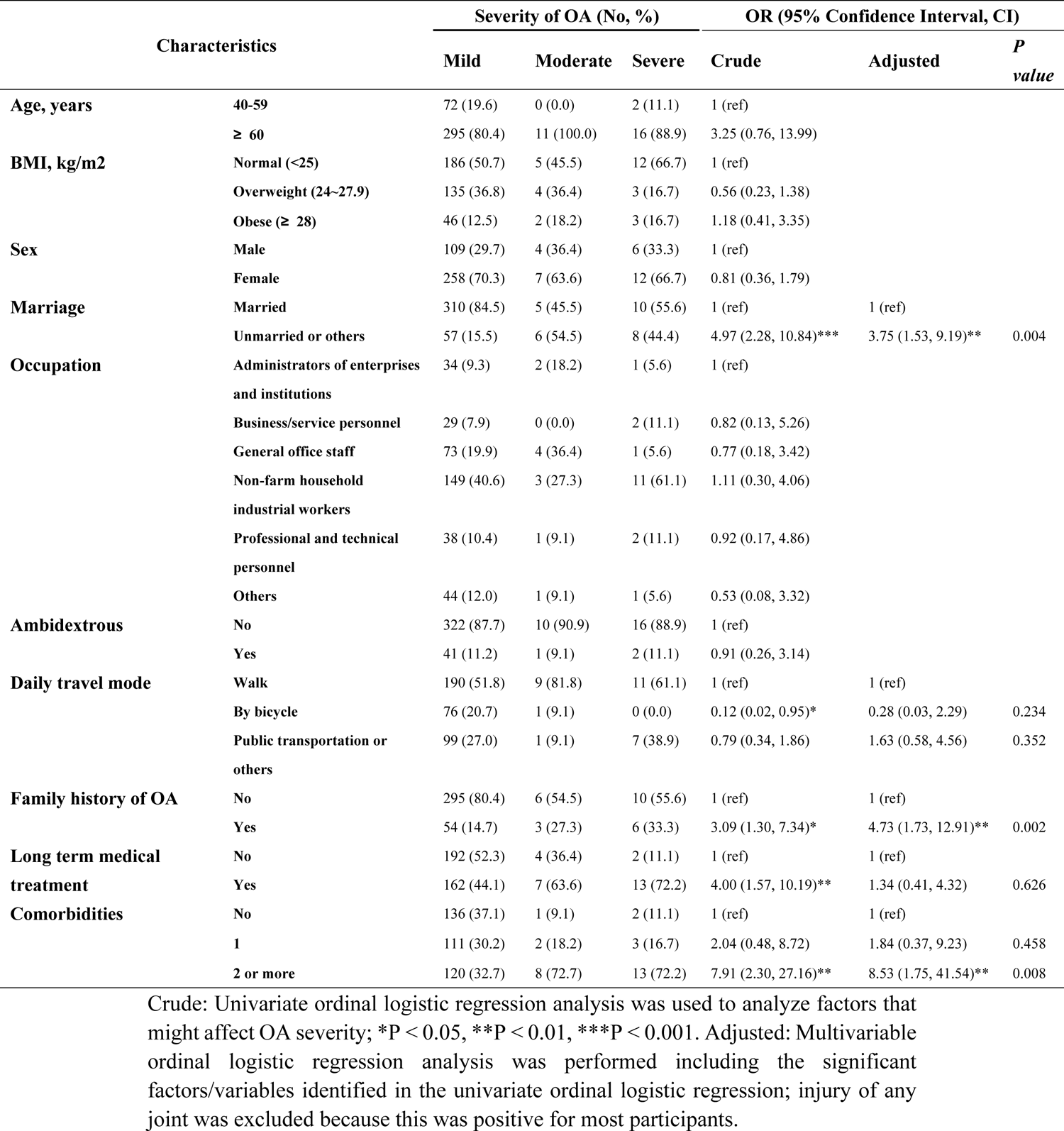
Analysis of the association of factors with OA severity.

## 4 Discussion

This study investigated the prevalence and factors associated with OA at any joint among those aged 40 and older. The knee OA was the most common among all region OA. The present study found that OA was associated with age ≥ 60, overweight BMI (24∼27.9 kg/m2), female sex, married status, a family history of OA, two or more comorbidities, and prior injury of any joint. Marriage status a family history of OA, and two or more comorbidities were found to be associated with OA severity.

Consistent with previous reports[17, 18], age and sex were significantly associated with OA in this study. Recent epidemiological studies have shown that the lack of estrogen after menopause may play a role in the pathogenesis of OA in elderly women, and that many women are more prone to OA after menopause[13, 19]. This suggests that sex hormones may contribute to the difference in OA between men and women. OA was also reported to be significantly affected by obesity[20], which is commonly believed to affect joints through biomechanical loading and metabolic inflammatory pathways[21]. An apparent dose-response relationship between obesity and OA was observed in this research. The adjusted OR for obese BMI (≥ 28 kg/m2) did not reach the level of significance, but this may reflect sparse-data bias due to the small number of participants with obese BMI. A correlation between joint hypermobility and OA has been reported in community populations[22, 23]; hypermobility is thought to exert greater biomechanical stresses on articular cartilage, potentially increasing the risk of OA. Consistent with this, we found that the ambidextrous tended to be protected from OA, as were individuals who mainly used bicycle/public transportation. We found that having a professional or technical occupation (aOR = 0.51; 0.29, 0.87, P = 0.010) was associated with a significantly lower prevalence of OA, whereas the prevalence peaked for non-farm-household industrial workers. These findings were similar to those of a population-based study indicating that physically demanding work is a major risk factor for OA [24]. Previous reports posited that joint injury (meniscal injuries, dislocations, fractures[25] and ACL tears[26]) may be a major risk factor for the development of knee OA[27, 28]. Our study confirmed that there was a significant association between prior joint injury and OA. This may reflect that direct trauma to the tissue can disrupt normal joint kinematics and cause altered load distribution within the joint[12].

Previous research found that OA has a significant genetic component[29]. The heritable component of OA is reported to be approximately 50%[30] and polygenic[31]. Our results showed that a family history of OA made significant contributions to both the presence and severity of OA. Eirik Weldingh reported that a family history contributes to OA and that maternal OA consistently increases the risk of offspring OA across different OA locations and severities[32]. Similar to the findings of a previous report[12], those suffering from two or more comorbidities (i.e., diabetes, cancer, lung disease, stroke, heart disease) were more likely to report OA, especially more severe OA. The relationship between OA and comorbidities, such as coronary heart disease (CHD), is not yet completely understood. It may partly reflect shared underlying disease mechanisms or risk factors and the influenced of shared treatments or those with effects on both diseases, such as NSAIDs[33]. There is a paucity of research on the relationship between marital status and OA, especially the severity of OA. A study in Taiwan revealed that higher disability could be explained by unmarried status among arthritic older patients[34], and another found that married patients had better overall outcomes after total knee arthroplasty [35]. Our present results suggest that married participants were less likely to have OA overall or severe OA than those unmarried or other, especially a milder OA. Being married has been generally found to confer benefits for one’s mental and physical health because it provides steady and reliable companionship, emotional intimacy, sexual partnership, and a buffer against ongoing stress[36, 37]. Notably, one-fifth of people with OA experience symptoms of depression and anxiety[38], almost half do not receive any mental healthcare support[39], and depression is related to worse pain, higher functional disability, and higher rates of mortality[40]. Thus, avoiding depression and anxiety may be an especially important domain to consider in the elderly population.

There are limitations to our analysis. First, reporting bias may be present if participants did not accurately recall their information (history of injury/OA, family history of OA, etc.). Second, our study was only descriptive and thus carries the possibility of residual confounding. In the future cohort, studies could be used to better demonstrate that the identified causes preceded the studied outcome. Although we stratified for major factors such as sex and BMI adjustment, this cross-sectional analysis did not exhaustively address other potential unmeasured confounders associated with OA, such as history of taking antihypertensive and anti-inflammatory drugs, which were not included in the questionnaire at the study-design stage. In the future, inflammatory factors such as C-reactive protein can be included to further analyze the mediating factors and the association between risk factors and OA. Third, despite our strenuous efforts to achieve a high response rate to the WOMAC questionnaire among participants with confirmed OA, the overall response rate to the WOMAC questionnaire was 71.9% (396/551), meaning that there may have been recruitment bias.

## 5 Conclusions

We found that confirmed OA in an elderly population was associated with a number of factors, including age ≥ 60, overweight, female sex, married status, professional and technical work, ambidextrous, not walking for daily travel, a family history of OA, two or more comorbidities, and prior injury of any joint. In addition, marriage status, a family history of OA, and comorbidity had significant contributions to the severity of OA. Appropriate prevention and treatment strategies, predominantly those targeting women and individuals who are older, unmarried, and have a family history of OA, should be developed to reduce the burden of the disease.

## Data Availability

Data cannot be shared publicly because of Ethics. Data are available on request from the corresponding author Yefeng Zhao(.)and Dr.Weibing Wang for researchers who meet the criteria for access to confidential data.

## Acknowledgments

We gratefully acknowledge all of the health care workers in Liqun hospital for Collection of questionnaires in this study.

## Authors’ contributions

XL and BZ contributed equally. YFZ, YW and WBW contributed equally to the correspondence work. XL, SFX and BZ analysed the data and drafted the manuscript. WBW and YFZ conducted critical revisions of the manuscript. All authors read and approved the final manuscript.

## Availability of data and materials

The data presented in this study are available on request from the corresponding author Yefeng Zhao(.)and Dr.Weibing Wang.

## Ethics approval and consent to participate

The study was approved by the Ethics Committee of the Shanghai Putuo Liqun Hospital (No. RT-201911).

## Consent for publication

Not applicable.

